# The impact of emerging *Plasmodium knowlesi* on accurate diagnosis by light microscopy: a systematic review and modelling analysis

**DOI:** 10.1101/2021.09.08.21263294

**Authors:** John H. Huber, Margaret Elliott, Cristian Koepfli, T. Alex Perkins

## Abstract

The five *Plasmodium* spp. that cause human malaria appear similar under light microscopy, which raises the possibility that misdiagnosis could routinely occur in clinical settings. Assessing the extent of misdiagnosis is of particular importance for monitoring *P. knowlesi*, which co-circulates with the other *Plasmodium* spp. We performed a systematic review and meta-analysis of studies comparing the performance of microscopy and PCR for diagnosing malaria in settings with co-circulation of the five *Plasmodium* spp. We assessed the extent to which co-circulation of *Plasmodium* parasites affects diagnostic outcomes. We fit a Bayesian hierarchical latent class model to estimate variation in microscopy sensitivity and specificity. Mean sensitivity of microscopy was low, yet highly variable across *Plasmodium* spp., ranging from 41.7% (95% CI: 22.8 – 64.1%) for *P. falciparum* and 40.3% (22.0 – 61.5%) for *P. vivax* to 0.119% (0.0121 – 0.640%) for *P. knowlesi*, 7.57% (2.66 – 22.0%) for *P. malariae*, and 0.180% (0.00491 – 1.21%) for *P. ovale*. Observed PCR prevalence was positively correlated with estimated microscopic sensitivity and negatively correlated with estimated microscopic specificity, though the strength of the associations varied by species. Our analysis suggests that co-circulation of *Plasmodium* spp. undermines the accuracy of microscopy. Sensitivity was considerably lower for *P. knowlesi, P. malariae*, and *P. ovale*. The negative association between specificity and prevalence imply that less frequently encountered species may be misdiagnosed as more frequently encountered species. Together, these results suggest that the burden of *P. knowlesi, P. malariae*, and *P. ovale* may be underappreciated in a clinical setting.

## INTRODUCTION

*Plasmodium falciparum* and *Plasmodium vivax* are the predominant parasites responsible for human malaria worldwide, causing an estimated 193.9 million and 14.3 million clinical cases in 2017, respectively^1,2^. Given their contribution to the global burden of malaria, *P. falciparum* and *P. vivax* remain the primary focus of public health efforts aimed at achieving malaria elimination and eradication^3^. However, three additional *Plasmodium* species—*P. knowlesi, P. malariae*, and *P. ovale*—also cause clinical episodes of malaria, though their overall burden remains uncertain due in part to routine misdiagnosis in a clinical setting^4^.

Diagnosis of clinical malaria is routinely performed using light microscopy (LM). In addition to challenges presented by low or submicroscopic parasite densities^5^, accurate speciation of malaria parasites using LM depends upon identification of morphological features characteristic of the *Plasmodium* spp. causing the infection. However, morphological similarities across the five *Plasmodium* spp., such as the resemblance of early blood-stage *P. knowlesi* parasites to *P. falciparum* parasites and late blood-stage *P. knowlesi* parasites to *P. malariae* parasites^3,6^, undermine species-level identification of *Plasmodium* infections^7^.

In settings with co-circulation of the *Plasmodium* spp., routine misdiagnosis by light microscopy has consequences for clinical decision-making and malaria control. The choice of therapeutic is specific to the *Plasmodium* spp. causing the infection^8^, so misdiagnosis may lead to ineffective treatment of blood-stage parasites for all *Plasmodium* spp. infections as well as ineffective or no treatment of liver-stage parasites for *P. vivax* and *P. ovale* infections.

Additionally, misdiagnosis may present challenges for monitoring *Plasmodium* spp. transmission, particularly for the emerging zoonosis *P. knowlesi*. Currently, the extent of human-to-human transmission of *P. knowlesi* is unknown^3,9^, and misdiagnosis could cause transmission clusters to appear smaller than their actual size, leading to an underestimate of transmission^10^.

Consequently, measuring the extent of misdiagnosis by light microscopy is of importance to public health, and past studies have directly compared the diagnostic performance of light microscopy to polymerase chain reaction (PCR) in settings with co-circulation of *Plasmodium* spp. parasites^7,11–22^. However, these studies varied in their criteria for enrollment and other study design features, as well as in the epidemiological settings in which they were conducted. This makes direct comparison of their estimates of diagnostic performance challenging.

To synthesize across these studies, we performed a systematic review and meta-analysis of the diagnostic performance of light microscopy in settings with co-circulation of all five *Plasmodium* spp. We identified studies that compared the diagnostic performance of light microscopy to PCR and fit a hierarchical Bayesian latent class model to estimate study-level and species-level sensitivities and specificities of light microscopy. Finally, to assess whether the perceived prevalences of the five species might affect diagnostic outcomes of light microscopy, we explored the relationship between the estimates of sensitivity and specificity that we obtained and the observed PCR prevalences in each study.

## METHODS

### Search Strategy and Selection Criteria

In this systematic review and meta-analysis, JHH searched MEDLINE and Web of Science for studies published in English before April 30^th^, 2020 that compared the diagnostic performance of light microscopy (LM) and polymerase chain reaction (PCR) for *Plasmodium* spp. infections. We identified studies using the following search strategy: (*Plasmodium* OR malaria) AND *knowlesi* AND ((microscopy OR microscopic OR blood film OR thick film OR thin film) AND (PCR or “polymerase chain reaction”)).

Studies met the criteria for inclusion if they compared the performance of LM and PCR on clinical samples collected in areas with endemic transmission of all five *Plasmodium* spp. that naturally infect humans (i.e., *P. falciparum, P. vivax, P. knowlesi, P. malariae*, and *P. ovale*). Furthermore, studies were only included if they used a PCR protocol that was capable of amplifying DNA from all five *Plasmodium* spp. Prior to 2004, the public health burden of *P. knowlesi* was underappreciated^6^, so restricting our search to studies published between 2004 and 2021 ensured that the diagnostic outcomes were representative of the current clinical and diagnostic landscape.

The data were extracted from each study that met the criteria for inclusion. Because the data in each study was anonymized and reported in aggregate, ethical approvals were not required according to a determination from the University of Notre Dame Institutional Review Board that this study did not constitute human subjects research.

### Data Analysis

#### Overview

To model the diagnostic performance of LM and PCR, we constructed a Bayesian hierarchical latent class model of clinical diagnosis^23^. In this model, the probability *p*_*ijkl*_ of obtaining LM outcome *i* ∈ {1,0} and PCR outcome *j* ∈ {1,0} for *Plasmodium* spp. *k* in study *l* is

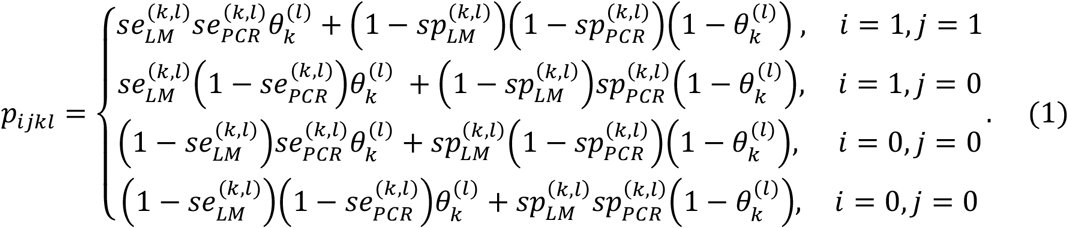

In eq. (1), 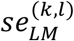 and 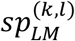 are the sensitivity and specificity of LM for *Plasmodium* spp. *k* in study 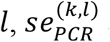 and 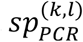 are the sensitivity and specificity of PCR for *Plasmodium* spp. *k* in study *l*, and 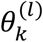 is the prevalence of *Plasmodium* spp. *k* among participants in study *l*. In many cases, 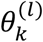 is not equivalent to community prevalence, due to characteristics of the study participants that are not representative of the surrounding community.

A key assumption of the latent class model is that diagnostic outcomes are conditionally independent. That is, by assuming that the sensitivity and specificity of a given diagnostic method is unique for each *Plasmodium* spp., the outcome that we observe for one parasite species using a given diagnostic method is modelled as independent of the outcomes observed for all other parasite species and diagnostic methods. This implies that the probability of a set of diagnostic outcomes observed across all five *Plasmodium* spp. is equal to the product of the probabilities of the diagnostic outcome for each *Plasmodium* spp. Nevertheless, the assumption of conditional independence still allows for interactions between *Plasmodium* spp. that arise in the context of misdiagnosis. For example, if a *P. knowlesi* infection is misdiagnosed as a *P. malariae* infection using LM, then the sensitivity of LM for *P. knowlesi* and the specificity of LM for *P. malariae* are penalized, because both a false-negative *P. knowlesi* diagnosis and a false-positive *P. malariae* diagnosis occurred.

#### Accounting for Differences in Study Design

Studies identified in the systematic review varied by enrollment and diagnostic criteria. Where enrollment in a study required a positive *Plasmodium* spp. diagnosis by LM or PCR, we normalized the probabilities of eq. (1), such that they were consistent with the subset of possible diagnostic outcomes that could be observed with that study design.

Because *P. knowlesi* and *P. malariae* share morphological similarities^3,6^, studies may diagnose samples as “*P. knowlesi* / *P. malariae*” with LM, indicating that the microscopist identified *P. knowlesi* and/or *P. malariae* parasites in the sample but could not make a more precise mono-infection or co-infection diagnosis. To accommodate this in our model, we defined parameters 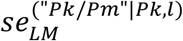 and 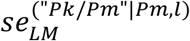 as the sensitivities of LM to diagnose samples in study *l* as “*P. knowlesi* / *P. malariae*”, given that the samples contained *P. knowlesi* and *P. malariae* parasites, respectively.

#### Inference

Using PCR as the gold standard, we fit a hierarchical latent class model of clinical diagnosis to the studies identified in our systematic review to estimate study-level and species-level variation in LM sensitivity and specificity for each *Plasmodium* spp. We fit our model in a Bayesian framework using Stan by running four independent chains of 2,000 samples, each with a warm-up period of 1,000 samples^24^. We tested for convergence with the Gelman-Rubin statistic and pooled the independent chains for a posterior distribution of 4,000 samples. Supplementary analyses were performed to evaluate the sensitivity of our posterior estimates to the inclusion of each study and to the assumption of PCR as the gold standard. See the Supporting Information for more details.

## RESULTS

Of the 176 unique studies identified using MEDLINE and Web of Science, 12 studies met the inclusion criteria, and their data comparing LM and PCR diagnostic performance were extracted (Fig. 1). The studies enrolled febrile patients presenting in health clinics in Malaysia (n = 7), Thailand (n = 2), Indonesia (n = 1), Myanmar (n = 1), and China (n = 1). Among the 12,690 total patients enrolled across the 12 studies, the pooled percentage positive by PCR was 17% for *P. falciparum*, 24% for *P. vivax*, 38% for *P. knowlesi*, 1.7% for *P. malariae*, and 0.36% for *P. ovale* (Table 1). The studies varied widely in the percentage positive by PCR across *Plasmodium* spp., encompassing a wide range of epidemiological settings and study designs (i.e., criteria for patient enrollment) in which we estimated LM diagnostic performance.

**Figure 1.**
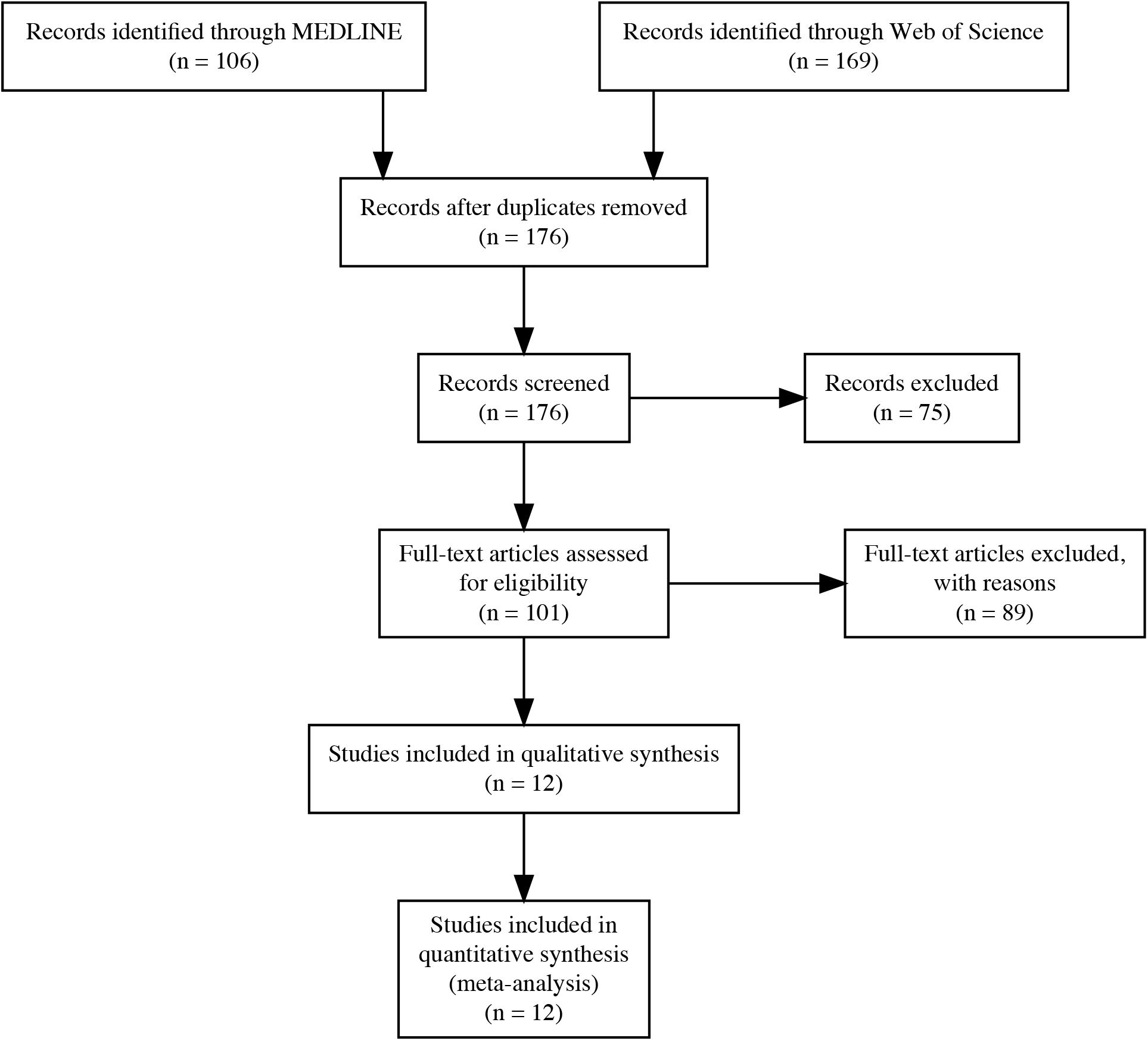
PRISMA Flow Diagram. Flow diagram of the studies identified using MEDLINE and Web of Science for the systematic review.

**Table 1.** Reported Plasmodium spp. percentage positive by diagnostic method. The percentage of study participants positive by LM and PCR are reported for each Plasmodium spp. in each study, along with the study location, year, and total number and type of samples collected. For Pk and Pm LM percentage of study participants positive, * indicates that the percentage positive was computed using a diagnosis of “Pk/Pm.”

| Ref. | Location | Samples | Type | Years | *P. falciparum* |  | *P. vivax* |  | *P. knowlesi* |  | *P. malariae* |  | *P. ovale* |  |
| --- | --- | --- | --- | --- | --- | --- | --- | --- | --- | --- | --- | --- | --- | --- |
|  |  |  |  |  | PCR | LM | PCR | LM | PCR | LM | PCR | LM | PCR | LM |
| 15 | Malaysia | 960 | LM+ | 2001- | 26 | 23 | 51 | 45 | 28 | 0.0 (0) | 0.42 | 33 | 0.73 | 0.21 |
|  |  |  |  | 2006 | (249) | (218) | (487) | (430) | (266) |  | (4) | (312) | (7) | (2) |
| 19 | Thailand | 1,874 | Any | 2006- | 41 | 37 | 64 | 54 | 0.53 | 0.0 (0) | 1.3 | 0.16 | 0.96 | 0.0 |
|  |  |  | Febrile | 2007 | (762) | (687) | (1192) | (1021) | (10) |  | (24) | (3) | (18) | (0) |
| 16 | Malaysia | 189 | LM+ | 2008- | 37 | 39 | 35 | 35 | 22 | 0.0 (0) | 1.1 | 27 | 0.0 | 0.0 |
|  |  |  |  | 2011 | (70) | (74) | (66) | (67) | (42) |  | (2) | (51) | (0) | (0) |
| 11 | Malaysia | 461 | LM+ | 2009- | 7.2 | 12 | 16 | 13 | 85 | 92* | 0.87 | 2* | 0.0 | 0.0 |
|  |  |  |  | 2011 | (33) | (54) | (74) | (59) | (392) | (425) | (4) | (425) | (0) | (0) |
| 13 | Malaysia | 227 | Any | 2008- | 42 | 41 | 41 | 38 | 16 | 0.0 (0) | 0.88 | 15 | 0.044 | 0.044 |
|  |  |  | Febrile | 2010 | (95) | (92) | (92) | (87) | (36) |  | (2) | (33) | (1) | (1) |
| 21 | China | 560 | Any | 2008- | 28 | 28 | 51 | 49 | 0.36 | 0.0 (0) | 1.6 | 0.0 (0) | 2.7 | 0.0 |
|  |  |  | Febrile | 2012 | (159) | (156) | (288) | (276) | (2) |  | (9) |  | (15) | (0) |
| 7 | Malaysia | 303 | PCR+/LM+ | 2010- | 43 | 47 | 16 | 16 | 44 | 42* | 0.66 | 42* | 0.0 | 0.0 |
|  |  |  |  | 2011 | (130) | (142) | (47) | (47) | (132) | (128) | (2) | (128) | (0) | (0) |
| 12 | Thailand | 297 | Any | 2012 | 9.1 | 3.4 | 8.8 | 5.1 | 0.0 (0) | 0.0 (0) | 0.34 | 0.34 | 0.0 | 0.0 |
|  |  |  | Febrile |  | (27) | (10) | (26) | (15) |  |  | (1) | (1) | (0) | (0) |
| 20 | Malaysia | 457 | LM+ | 2012- | 11 | 12 | 32 | 30 | 58 | 40 | 0.22 | 18 | 0.044 | 0.022 |
|  |  |  |  | 2013 | (51) | (54) | (144) | (138) | (267) | (185) | (1) | (81) | (2) | (1) |
| 17 | Myanmar | 90 | Any | 2013- | 44 | 43 | 20 | 19 | 0.0 (0) | 0.0 (0) | 1.1 | 1.1 (1) | 0.0 | 0.0 |
|  |  |  | Febrile | 2015 | (40) | (39) | (18) | (17) |  |  | (1) |  | (0) | (0) |
| 18 | Indonesia |  | Any | 2015 |  |  |  |  |  |  |  |  |  |  |
|  |  |  | Febrile |  |  |  |  |  |  |  |  |  |  |  |
|  | Batubara | 1,270 |  |  | 6.2 | 3.4 | 10 | 6.5 | 12 | 0.0 (0) | 1.2 | 0.0 (0) | 0.0 | 0.0 |
|  |  |  |  |  | (79) | (43) | (131) | (83) | (148) |  | (15) |  | (0) | (0) |
|  | Langkat | 544 |  |  | 16 | 9.6 | 16 | 10 | 5.9 | 0.0 (0) | 1.1 | 0.18 | 0.0 | 0.0 |
|  |  |  |  |  | (87) | (52) | (85) | (56) | (32) |  | (6) | (1) | (0) | (0) |
|  | South Nias | 1,917 |  |  | 12 | 15 | 17 | 6.8 | 14 | 0.0 (0) | 5.6 | 0.0 (0) | 0.0 | 0.0 |
|  |  |  |  |  | (227) | (294) | (321) | (131) | (263) |  | (107) |  | (0) | (0) |
| <sup>14</sup> | Malaysia | 3,541 | PCR+/LM+ | 2015- | 4.9 | 4.9 | 2.5 | 2.5 | 92 | 93* | 1.1 | 93* | 0.085 | 0.0 |
|  |  |  |  | 2017 | (173) | (174) | (87) | (87) | (3255) | (3280) | (40) | (3280) | (3) | (0) |
| **Total** |  | 12,690 |  |  | 17 | 16 | 24 | 20 | 38 | 32 | 1.7 | 34 | 0.36 | 0.032 |
|  |  |  |  |  | (2182) | (2089) | (3058) | (2514) | (4845) | (4018) | (218) | (4316) | (46) | (4) |

After validating on 200 simulated datasets (Fig. S1), we applied our hierarchical latent class model to the data from the 12 studies identified in the systematic review. Our fitted hierarchical latent class model captured the variation in the observed data (Fig. S2) and revealed differences in LM diagnostic performance across *Plasmodium* spp. (Fig. 2). Specifically, we estimated that the species-level mean sensitivity of LM was 41.7% (95% CI: 22.8 – 64.1%) for *P. falciparum*, 40.3% (22.0 – 61.5%) for *P. vivax*, 0.119% (0.0121 – 0.640%) for *P. knowlesi*, 7.57% (2.66 – 22.0%) for *P. malariae*, and 0.180% (0.00491 – 1.21%) for *P. ovale*. Species-level mean specificity of LM was consistently high across *Plasmodium* spp. and was estimated at 98.5% (96.7 – 99.4%) for *P. falciparum*, 98.8% (97.3 – 99.5%) for *P. vivax*, 100% (99.9 – 100%) for *P. knowlesi*, 99.3% (98.0 – 99.8%) for *P. malariae*, and 100% (100 – 100%) for *P. ovale*. These results were robust to the assumption of PCR as the gold standard (Fig. S4).

**Figure 2.**
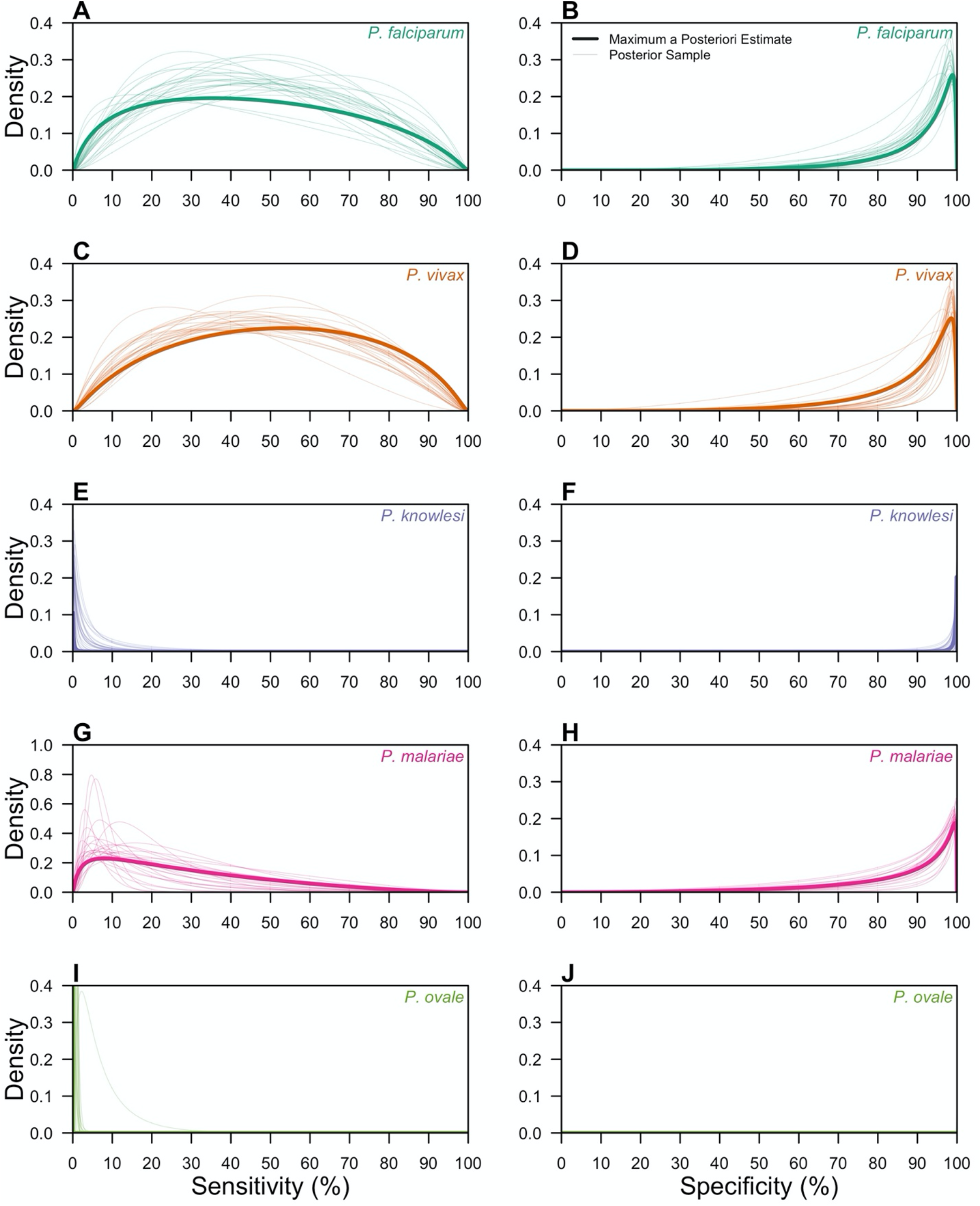
Group-level posterior estimates of LM diagnostic performance. The group-level distributions of LM sensitivity (A, C, E, G, I) and LM specificity (B, D, F, H, I) are shown for Plasmodium falciparum (teal; A, B), Plasmodium vivax (orange; C, D), Plasmodium knowlesi (purple; E, F), Plasmodium malariae (pink; G, H), and Plasmodium ovale (green; I, J). Thick lines are the maximum a posteriori estimates, and thin lines are 25 posterior samples.

The variance in the hierarchical distributions of LM diagnostic performance in Fig. 2 revealed the extent of variation across studies, a feature that depended upon both the diagnostic measure and the *Plasmodium* spp. considered (Fig. 3). In general, for a given *Plasmodium* spp., we estimated greater variation across studies in LM sensitivity than specificity. For example, for *P. falciparum*, we estimated that logit-transformed standard deviation of LM sensitivity was 1.61 (1.26 – 2.09), suggesting that the range of sensitivities that fell within one standard deviation of the mean was 12.5 – 78.2%. By comparison, the estimated logit-transformed standard deviation of LM specificity for *P. falciparum* was 1.48 (1.13 – 1.93), corresponding to a range of specificities within one standard deviation of the mean of 93.8 – 99.7%. Across *Plasmodium* spp., study-level estimates of LM sensitivity and specificity for *P. falciparum, P. vivax*, and *P. malariae* exhibited greater variation than for *P. knowlesi* and *P. ovale*.

**Figure 3.**
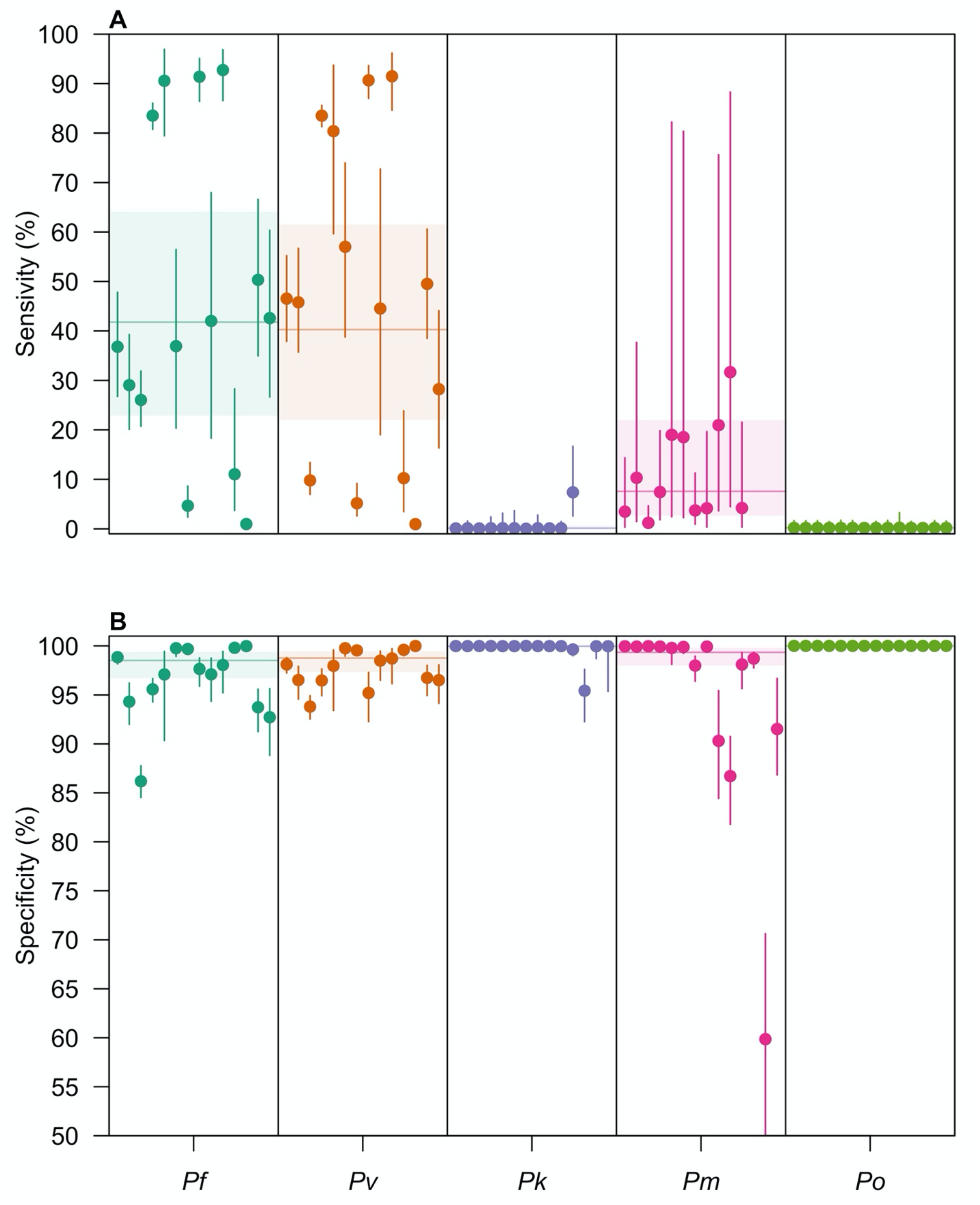
Site-level posterior estimates of LM diagnostic performance. The site-level posterior estimates of (A) LM sensitivity and (B) LM specificity are shown for P. falciparum (teal), P. vivax (orange), P. knowlesi (purple), P. malariae (pink), and P. ovale (green). Circles are the median posterior estimate, and the vertical segment is the 95% credible interval. The horizontal line is the posterior median of the group-level mean, and the horizontal shaded region is the corresponding 95% credible interval.

Because the estimates of LM diagnostic performance varied across the unique studies, we then assessed its relationship with observed PCR prevalence (Fig. 4). We observed strong, positive correlations between PCR prevalence and LM sensitivity for *P. falciparum* (Pearson Correlation Test: ρ = 0.66; p = 0.01) and *P. knowlesi* (ρ = 0.83; p = 0.0015) and weaker, positive correlations for *P. vivax* (ρ = 0.40) and *P. ovale* (ρ = 0.012), though the latter two were not statistically significant associations. By contrast, for *P. malariae*, there was a negative correlation between PCR prevalence and LM sensitivity, though this estimate was also not statistically significant. Across *Plasmodium* spp., increased PCR prevalence of *P. knowlesi* was associated with reduced sensitivity of LM for *P. vivax* (ρ = -0.58; p = 0.029), highlighting a possible source of misdiagnosis. Additional negative associations were observed between PCR prevalence and LM sensitivity across *P. falciparum, P. knowlesi*, and *P. malariae*—*Plasmodium* spp. that are commonly misdiagnosed—though these quantities were not statistically significant.

**Figure 4.**
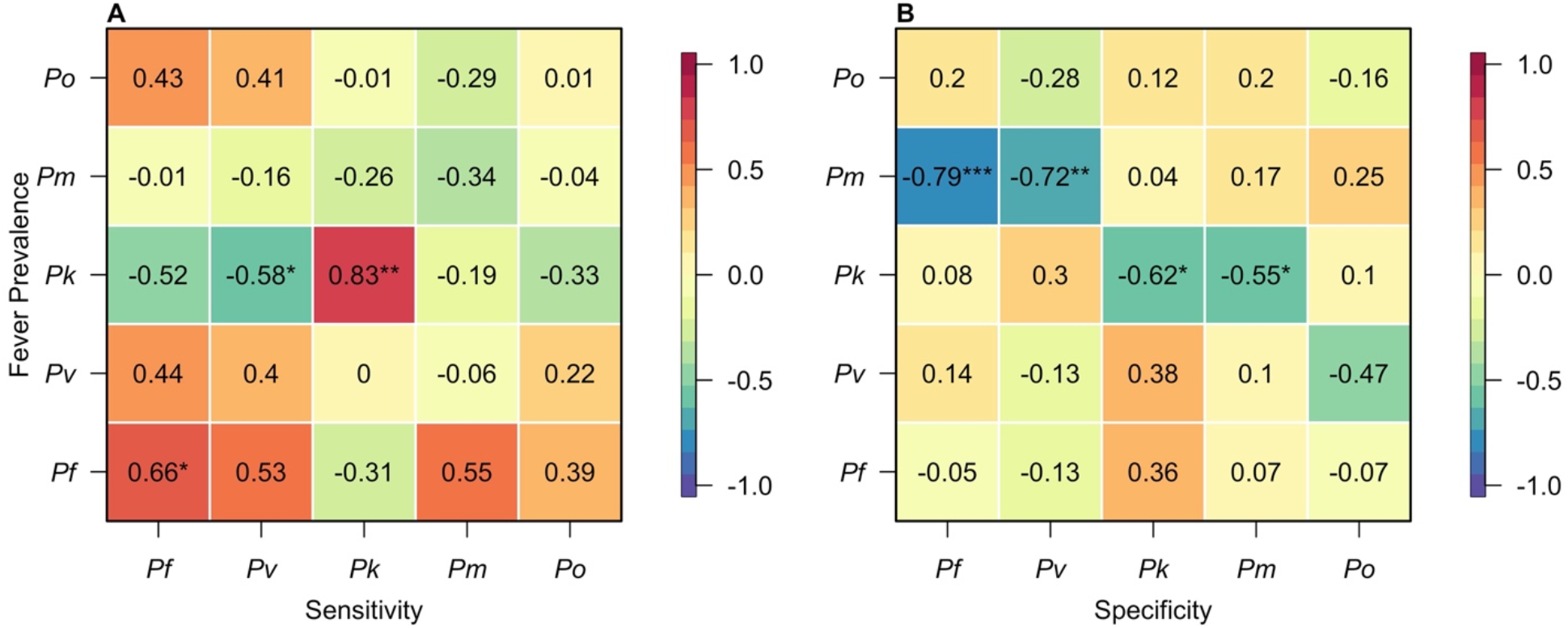
Correlations between Plasmodium spp. fever prevalence and LM diagnostic performance. The correlations between Plasmodium spp. fever prevalence and (A) LM sensitivity and (B) LM specificity are reported. The color represents the strength of the correlation, with red colors denoting positive correlations and blue colors denoting negative correlations. ‘*’ denotes p < 0.05, ‘**’ denotes p < 0.01, and ‘***’ denotes p < 0.001.

Generally, for a given *Plasmodium* spp., PCR prevalence was negatively associated with LM specificity (Fig. 4B). For *P. knowlesi*, this association was strong and statistically significant (ρ = -0.62; p = 0.017), and additional weaker, yet statistically non-significant relationships were observed for *P. falciparum, P. vivax*, and *P. ovale*. The opposite relationship was observed for *P. malariae*, though this quantity was not statistically significant. Across *Plasmodium* spp., increased prevalence of *P. malariae* was associated with reduced specificities of LM for *P. falciparum* (ρ = -0.79; p < 0.001) and *P. vivax* (ρ = -0.72; p = 0.0036), and increased prevalence of *P. knowlesi* was associated with reduced specificity of LM for *P. malariae*.

## DISCUSSION

In this systematic review and meta-analysis, we estimated study-level and species-level variation in the malaria diagnostic performance of light microscopy in epidemiological settings with co-circulation of the five *Plasmodium* spp. that cause human malaria. Our analysis revealed consistently low sensitivity of light microscopy to diagnose *Plasmodium* spp. infection. Sensitivity of light microscopy was estimated to be considerably lower for *P. knowlesi, P. malariae*, and *P. ovale* than for *P. falciparum* and *P. vivax*. This suggests that the burden of *P. knowlesi, P. malariae*, and *P. ovale* in a clinical setting may be appreciably underestimated, consistent with a recent study of *P. ovale* in health clinics in Kenya^4^.

Post-hoc analyses revealed relationships between the diagnostic performance of light microscopy and the epidemiological setting. In general, we noted a positive association between the PCR prevalence of a *Plasmodium* spp. and the sensitivity of light microscopy for that *Plasmodium* spp. This supports a phenomenon whereby the more a microscopist encounters samples of a *Plasmodium* spp. infection in a clinical setting, the more likely they are to accurately identify the *Plasmodium* spp. causing that infection. By contrast, increased PCR prevalence of a *Plasmodium* spp. was associated with high false-positive probabilities for that *Plasmodium* spp. This tendency to misdiagnose malaria infections as what is most commonly encountered in that clinical setting could lead to ineffective monitoring of less commonly encountered *Plasmodium* spp., such as *P. knowlesi, P. malariae, and P. ovale*.

The high probabilities of misdiagnosis estimated in this analysis could have important consequences for malaria treatment and control. Specifically, we observed an association between increased prevalence of *P. knowlesi* and reduced sensitivity of LM for *P. vivax*. This suggests that in areas with *P. knowlesi*, infections of *P. vivax* may not be correctly diagnosed and therefore may not receive radical cure therapy to clear hypnozoites and prevent relapse^25^. Additionally, increased prevalence of *P. malariae* was associated with reduced specificity of LM for *P. vivax*, indicating that many *P. malariae* infections may be misdiagnosed as *P. vivax*. This form of misdiagnosis could unnecessarily subject individuals to radical cure therapy, potentially increasing the risk of acute hemolysis among individuals deficient of the glucose-6-phosphate dehydrogenase enzyme^26^.

Routine diagnosis of clinical malaria by light microscopy may prevent effective monitoring of *P. knowlesi* transmission and burden. We estimated very low sensitivity of LM to diagnose *P. knowlesi* infections, given the morphological similarities of the parasites to those of *P. falciparum* and *P. malariae*^3,6^ as well as likely delays in familiarity of *P. knowlesi* among microscopists in the health systems. Sensitivity of LM was improved in settings that allowed for a “*P. knowlesi/P. malariae*” diagnosis, indicating that further speciation is often not possible. Although in practice *P. knowlesi* and *P. malariae* infections diagnosed by light microscopy are confirmed by PCR in many settings^11^, *P. knowlesi* infections misdiagnosed as *P. falciparum*, for instance, would not be subject to confirmatory PCR. This could lead to an underestimate of *P. knowlesi* burden, affecting epidemiological assessments of *P. knowlesi* infection risk.

Our analysis is subject to a number of limitations. Our systematic review may not have identified all studies with relevant data, including those studies not indexed by MEDLINE and Web of Science. We limited our review to studies that tested for all five *Plasmodium* spp. However, studies that included subsets of the five *Plasmodium* spp. could still be informative of species-level diagnostic performance. Our latent class model assumed conditional independence of diagnosis across *Plasmodium* spp. Future extensions of this work could account for the correlation structure of LM diagnosis across the *Plasmodium* spp., due to shared morphologies of the parasites. Additionally, to address non-identifiability issues, we assumed PCR to be the gold standard. Although sensitivity analyses revealed that our species-level estimates were robust to this assumption, future work could jointly estimate the diagnostic performance of both methods, providing a more complete characterization of misdiagnosis of *Plasmodium* spp. in settings with co-circulation of multiple *Plasmodium* species.

## Data Availability

All code and data to reproduce the analyses can be accessed at https://github.com/johnhhuber/malaria_misdiagonsis.git.

## NOTES

## Acknowledgments

The authors thank Parker Ladwig for helpful advice on conducting a systematic review.

## Author Contributions

J.H.H. and T.A.P. conceived of the study. J.H.H. developed the methodology and performed the systematic review and analyses. J.H.H. and M.E. verified the underlying data. J.H.H., M.E., C.K., and T.A.P. interpreted the results. J.H.H. and T.A.P. wrote the initial draft, and all authors provided comments on the final draft. All authors had access to the underlying data.

## Disclaimer

The funding sources had no role in the study design, data collection, analysis, interpretation, or writing of the report.

